# Knowledge, Attitude, and Practices, Hand Hygiene, and Preferred Food Safety Interventions of Butchers Operating in Nairobi County, Kenya

**DOI:** 10.1101/2025.10.10.25337753

**Authors:** Winnie Aketch Ogutu, Peter Omemo, Patricia Koech, Maureen Kuboka, Linnet Ochieng, Max Korir, Arshnee Moodley, Florence Mutua, Delia Grace

## Abstract

Foodborne diseases are a major public health concern globally, often linked to unhygienic food handling. In Kenya, meat is frequently sold through small, informal butcher shops with limited infrastructure. This study assessed the food safety knowledge, attitudes, and practices (KAP) of butchers, evaluated microbial contamination of their hands, explored latent dimensions of KAP, and food safety interventions preference. We conducted a cross-sectional study among 200 butchers randomly selected from four peri-urban areas of Nairobi. A semi-structured questionnaire captured KAP and intervention preferences, with pre- and post-meat handling hand contamination assessed using contact plate cultures. Psychometric properties of KAP domains were assessed using principal component analysis (PCA), Cronbach’s alpha, and item response theory (IRT). Exploratory factor analysis (EFA) and cluster analysis were used to segment butchers by intervention preferences. Respondents were predominantly male (96.9%), 42.7% were aged 20–30 and 52% reported > 5 years’ work experience. Knowledge of regulatory requirements was high (98% were aware that meat must be inspected), yet awareness of WHO’s Five Keys was extremely low (1%). Attitudes toward hygiene were generally positive, and many reported good practices (daily changing of towels (82%)). However, risky behaviors were common, including storing meat at ambient temperatures and selling ready-to-eat food alongside raw meat. Hand contamination exceeded acceptable limits in 64% of butchers before and 86% after meat handling. Psychometric analysis confirmed the validity of the KAP indices, with modest internal consistency and good discrimination for low-performing respondents. Cluster analysis revealed three butcher typologies: Collectivist Effective Oversight Avoiders (48%); Formal System-Embracers (37%); and Forward-Facing Self-Reliant (15%). Despite positive attitudes and good self-reported practices, high levels of contamination and risky behaviors persisted. Effective policy approaches should combine conventional training, regulation and infrastructure upgrading with innovative, demand-driven and technology-based interventions. Initial efforts could focus on the minority already receptive to consumer-focused solutions, using them as entry points for more modern and impactful approaches.

## Introduction

Safe food is essential for health, food security, livelihoods, economic development, and trade. Foodborne diseases (FBDs) present significant challenges to public health, especially in low and middle-income countries (LMICs), where infrastructure, institutions, education, awareness, and incentives to ensure safe food are inadequate. A conservatively estimated 600 million people fall ill annually because of consuming contaminated food, and 420,000 died in 2010 [1]. Most of this burden occurs in LMICs and Africa has the highest per capita burden [2].

While animal-source foods are a good source of nutrients [3], they are prone to contamination and are common causes of FBD [4]. Meat is an excellent medium for microbial survival and growth, due to its high-water content, low carbohydrate, and moderate pH [5]. Microbial contamination of raw beef meat can occur at any point along the meat value chain (slaughter, transport, preparation, and storage), but retail is an important point where contamination can be introduced or increase [6]. In many countries in eastern and southern Africa meat is mainly sold from butcher shops. Butchers purchase carcasses from slaughter operations and process them into meat cuts: some produce sausages, and it is increasingly common to sell ready-cooked meat to consumers who either take away or can sit and eat in small adjacent restaurants [7]. Butcher owners often employ staff to assist, and in the manuscript, these will be referred to as butchers.

Correct knowledge on food safety, positive attitude, and good practices such as hygienic handling of food can prevent contamination [8] and improve meat safety. However, there is limited data on knowledge, attitude and practice (KAP) of butchers in East Africa and no recent studies on bacterial contamination of butchers’ hands or its relationship with KAP.

As more evidence emerges on the health and economic burden of foodborne disease [9], so has the urgency of identifying interventions that can be effective in the informal food sector of LMICs, responsible for most of the burden [10]. A recent systematic literature review pointed to the need for interventions to go beyond training and information, to address behaviour and institutions [11], and as such, recent years have seen innovative approaches including mass-media campaigns, upskilling of informal workers and leveraging consumer demand for food safety [12,13, 14]. However, although key to the success of any intervention, there is little information on the attitude of informal sector workers to novel approaches based on behavior and incentives rather than training and awareness raising.

Therefore, this study was designed to assess KAP of butchers, determine bacterial contamination of the butchers’ hands, and understand their preferences regarding potential food safety interventions. The study was part of a larger project on improving the safety of meat sold in peri-urban areas of Nairobi. Part of the baseline results have been published by [15]. The findings were considered in the development of an intervention to improve meat safety in Nairobi County. Additionally, the findings are relevant to implementors, donors, academics, and everyone interested in improving food safety in LMICs.

## Materials and Methods

### Study area

This study was conducted in Nairobi County (01°17’11” S 36 °49’02” E) which hosts the capital of Kenya. Nairobi has a population of 4,397,073 people [16]. The County has a total of eleven sub counties. Four peri-urban areas were considered for this study: Kangemi, Huruma, Kawangware, and Waithaka (Figure 1). These sites were purposively selected based on their high concentration of butchery outlets and because this is where most residents in the category of low and middle-income class live [17].

**Figure 1:**
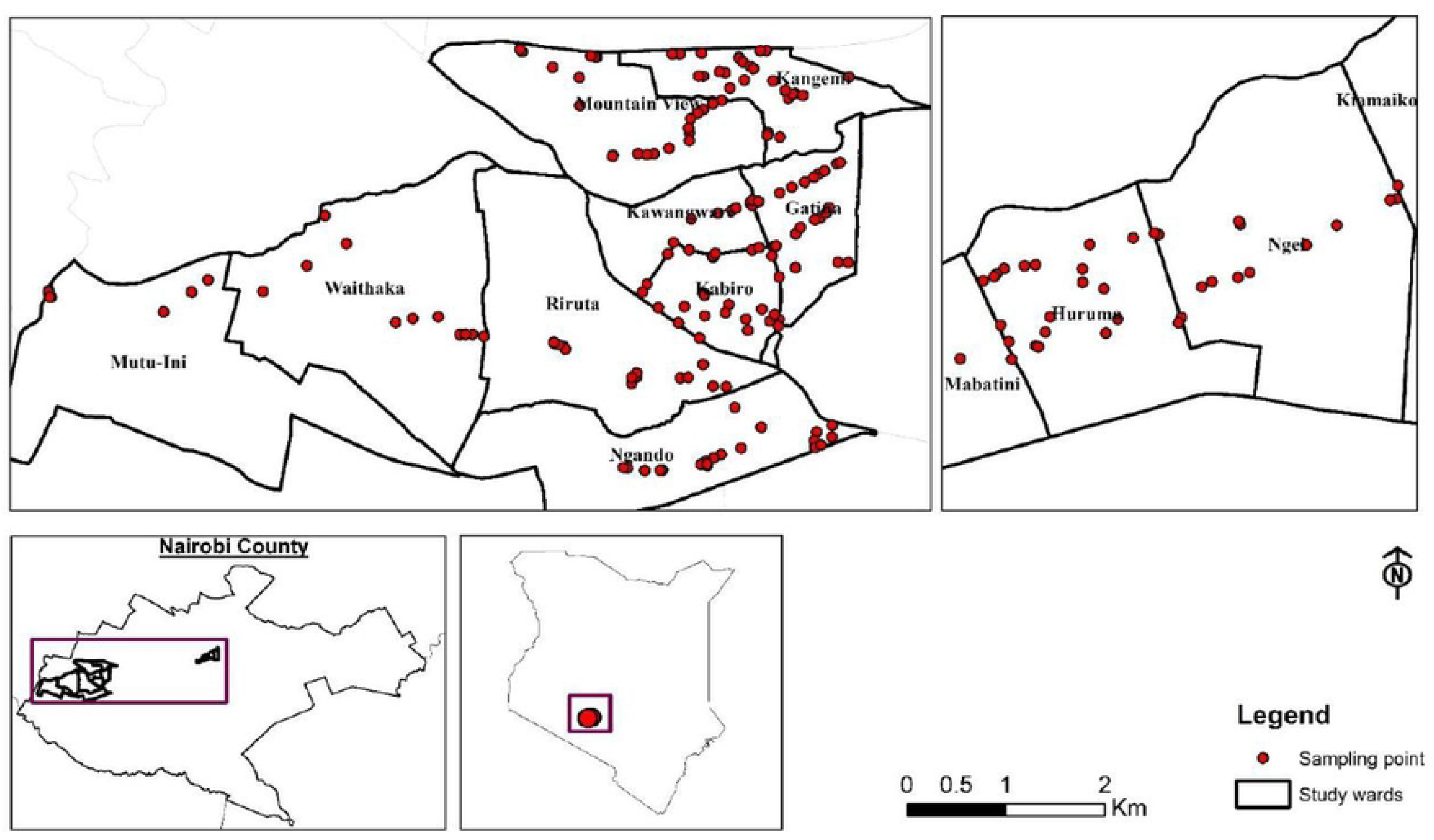
The four study areas: Kawangware, Kangemi, Huruma, and Waithaka.

### Sampling and sample size determination

Between February and April 2022, a census and GPS mapping of butcheries selling beef in the four study areas found 430 shops. The cross-sectional study served as a baseline for an intervention, with the sample size determined to detect a 15% reduction in unacceptable coliforms levels in the intervention group. This calculation resulted in a required sample of 340 butcheries, which was adjusted for finite population size to yield a final sample size of 189. To account for potential withdrawals or loss to follow-up, a total of 200 butcheries were included in the study.

### Data collection

A semi-structured questionnaire was developed, pretested, and the final version was translated into an Open Data Toolkit (ODK). The questionnaire had seven sections, namely, demographics, butcher shop details, knowledge, attitude, and practices, the most and least liked food safety interventions. The knowledge and attitude sections consisted of close-ended questions with three response options: agree, disagree, and don’t know. The practice questions were also close-ended, offering three choices: always, sometimes, and rarely. Questions were adapted from instruments used in previous KAP studies [18, 19].

For interventions, the most conventional were a government-led food safety and meat promotion campaign and formation of cooperative groups for butchers. Interventions widely used in high income countries and currently being advocated in Africa included a tamper-proof traceability label showing farm and abattoir of origin, a certification scheme whereby trained butchers would receive a certificate. Three novel consumer-demand led approaches were suggested: a brand label indicating high quality, safe meat; weekly testing of meat by trusted third parties with publication of results; and a hand-held device which consumers and butchers could use to test the freshness and safety of meat.

The 200 butcheries were randomly selected using the randomization function (RAND ()) in Microsoft Excel. The interviews were conducted from 18/05/2022 to 16/10/2022. Each selected shop was visited, and one person interviewed, either the owner of the butcher shop or the butcher, depending on who was available at the time of the study visit. The person interviewed was directly involved in the handling and selling of the meat. A written informed consent was obtained prior to administering the semi-structured questionnaire.

### Sample collection

Handprint samples were collected directly using sterile, pre-prepared nutrient agar plates. Two handprint samples were collected from each participant: one from the right hand and one from the left. The first handprint was collected before meat handling/selling, and the second after the meat was handled. Participants were instructed to gently press their hand onto the nutrient agar plate, for approximately five seconds. The plates were then packed and transported aseptically to the lab at 4℃ where they were incubated at 37℃ for 18 - 24 hours, after which they were assessed for microbial growth.

### Data management and analysis

The ODK data was downloaded as a Microsoft Excel file, cleaned, and subsequently exported to IBM SPSS (version 25) and Stata (version 17) for statistical analysis. Appropriate descriptive statistics were computed (tables, graphs). For each KAP question, frequency counts and percentages were calculated. We evaluated the psychometric properties of the KAP indices using a combination of classical and modern test theory. For the knowledge scale, principal component analysis (PCA) was performed on dichotomous items to assess dimensionality. A one-factor solution was retained based on eigenvalues >1 and item loading patterns, and internal consistency was assessed using Cronbach’s alpha. For the attitude and practice domains, which consisted of Likert-scale items, graded response models were fitted using two-parameter logistic response theory (IRT). Items with low discrimination (<0.5) or poorly ordered thresholds were considered for removal. Latent trait estimates (theta scores) were derived for each construct and used in downstream analyses to explore associations between KAP and other variables and for exploratory factor analysis. The Chi square and t-test were used to assess associations between variables. To explore underlying patterns in butcher preferences for food safety interventions, we conducted a principal factor analysis using principal factor extraction with orthogonal (varimax) rotation. The number of factors to retain was guided by the Kaiser criterion (eigenvalues >1) and the interpretability of the resulting factor structure.

To identify distinct typologies of butchers based on their intervention preferences, we conducted hierarchical cluster analysis using Ward’s linkage method on the three factor scores derived from the EFA. Ward’s method was selected because it minimizes within-cluster variance and tends to produce compact, similarly sized clusters appropriate for behavioural typologies. The optimal number of clusters was determined using the Calinski-Harabasz pseudo-F statistic and dendrogram inspection. Cluster validity was assessed through analysis of variance (ANOVA) to test whether clusters differed significantly on factor scores, and cross-validation was performed by examining associations between cluster membership and original intervention preferences using chi-square tests.

Bacterial growth was classified into three categories based on the number of colony-forming units (CFU) observed: low growth (0-49 colonies), medium growth (50-99 colonies), and heavy growth (100 or more colonies). The threshold of 100 CFU/cm^2^ was selected in line with South Africa’s Government Regulation 962 of 2012, issued under the Foodstuffs, Cosmetics and Disinfectants Act (No. 54 of 1972). This regulation permits a maximum of 100 viable organisms per cm^2^ after cleaning and sanitizing food contact surfaces. For this study, the same standard was applied to the hands of butchers, as their hands directly contact meat during handling, processing, and storage. contamination. A statistical significance level of 0.05 was used for all tests.

### Ethical Approvals

This study was approved by the International Livestock Research Institute Review and Ethics Committee (IREC); ILRI-IREC2021-62), Nairobi Metropolitan Services Ethic Review Board; REF: EOP/NMS/HS/132, National Commission for Science, Technology, and Innovation (NACOSTI-NACOSTI/P/22/15711; NACOSTI/P/24/39030), and Maseno University Scientific Ethics and Review Committee (MUSERC); MUSERC/01268/23. A written informed consent was obtained from each study participant.

## Results

### Demographic characteristics of the butchers

A total of 200 butchers participated in this study, however, data from only 192 participants were included in the analyses as eight responses were not saved in ODK during data collection. The socio-demographic characteristics of the participants are summarized in Table 1. Most respondents were male (96.9%) and aged between 20 −30 years (42.7%). Half had completed secondary education (50%), and just over half (51.6%) had >5 years of experience in selling meat. Additionally, 76% reported no prior training in food safety or meat handling, and only 25.5% possessed a valid medical health certificate. Almost all butchers (99%) were unfamiliar with the WHO’s *Five Keys to Food Safety* and Hazard Analysis Critical Control Point (HACCP) system.

**Table 1:**
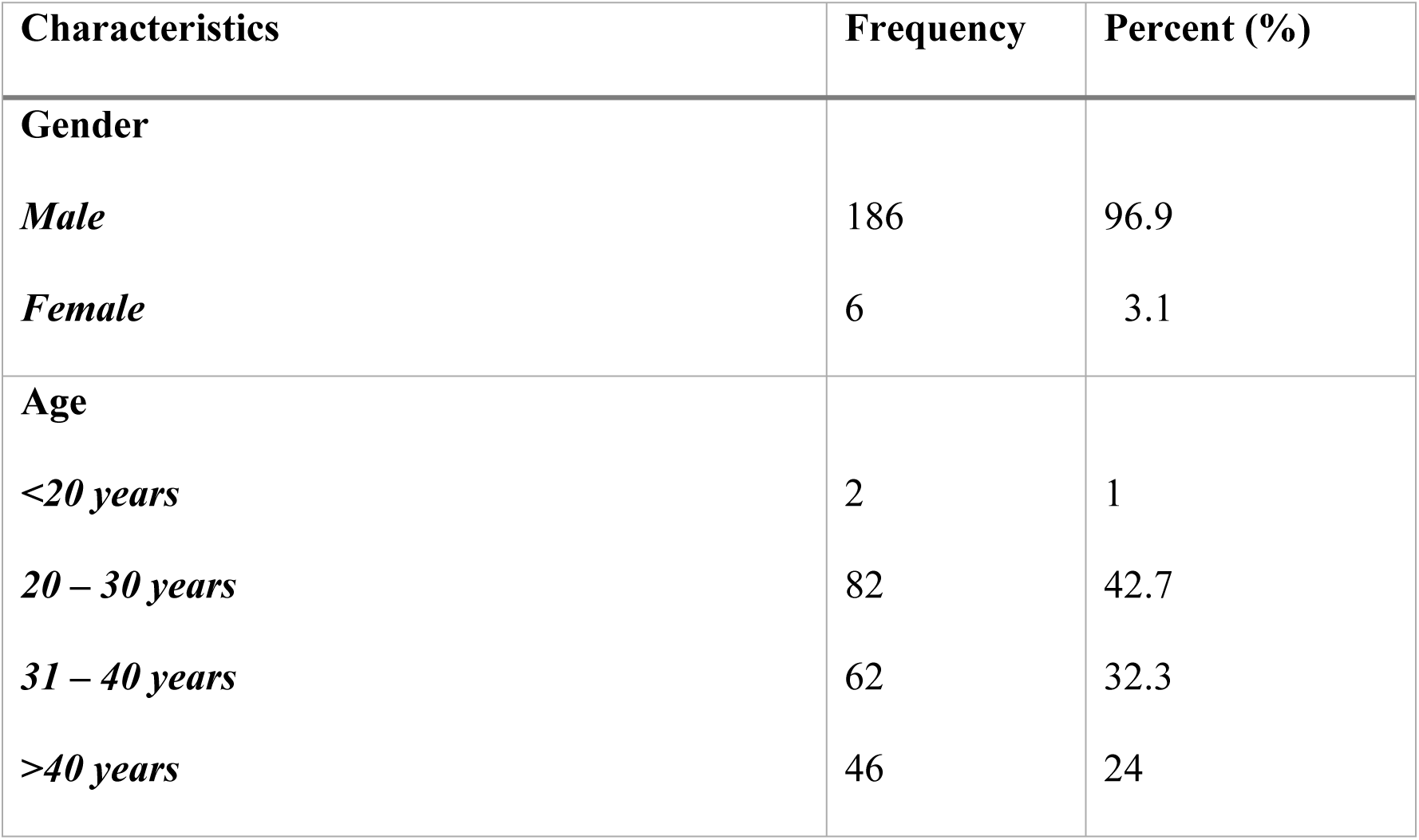
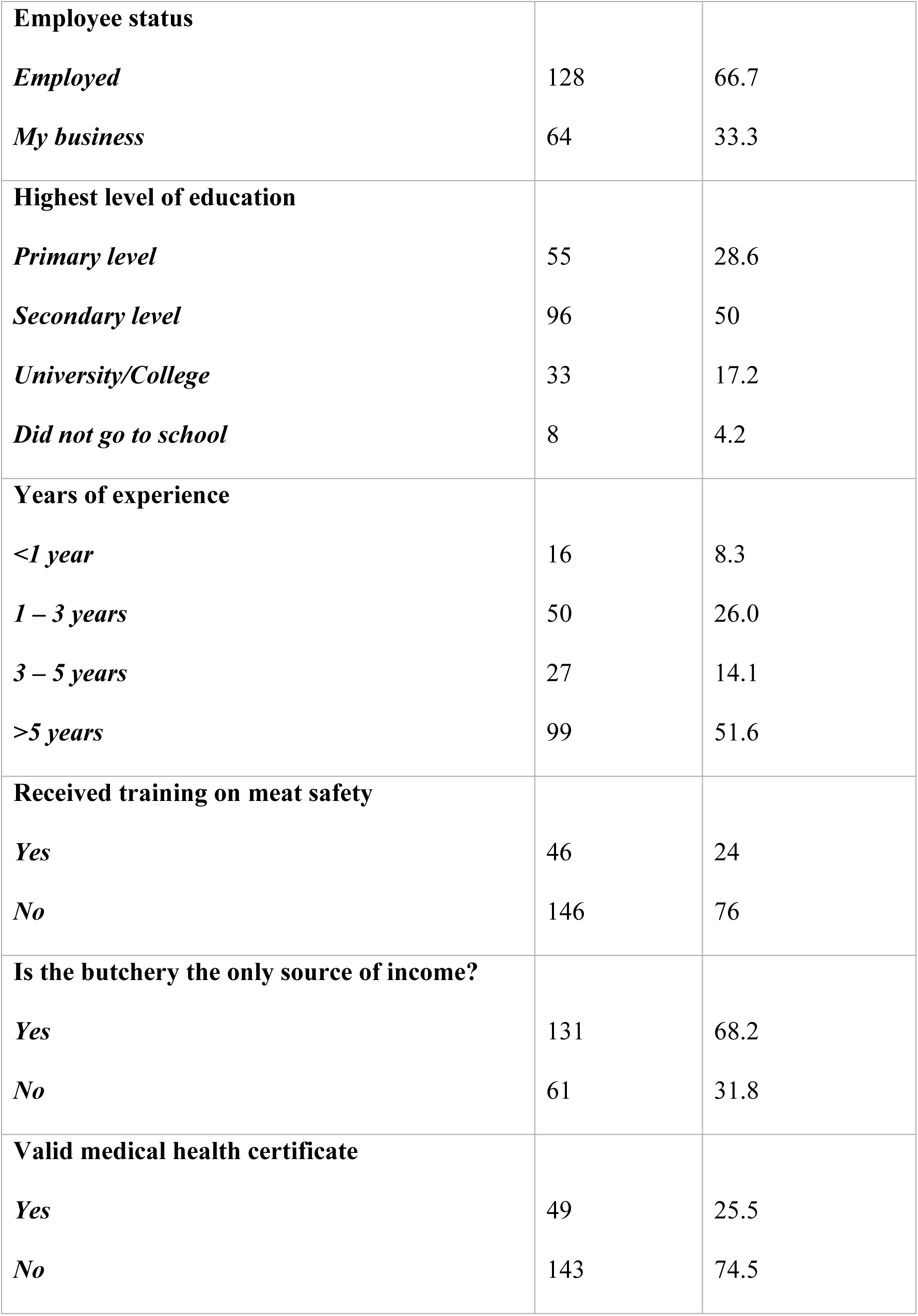
Demographic characteristics of butchers in the meat safety study, (May to October 2022)

| Characteristics | Frequency | Percent (%) |
| --- | --- | --- |
| **Gender** |  |  |
| *Male* | 186 | 96.9 |
| *Female* | 6 | 3.1 |
| **Age** |  |  |
| *<20 years* | 2 | 1 |
| *20 – 30 years* | 82 | 42.7 |
| *31 – 40 years* | 62 | 32.3 |
| *>40 years* | 46 | 24 |
| **Employee status** |  |  |
| *Employed* | 128 | 66.7 |
| *My business* | 64 | 33.3 |
| **Highest level of education** |  |  |
| *Primary level* | 55 | 28.6 |
| *Secondary level* | 96 | 50 |
| *University/College* | 33 | 17.2 |
| *Did not go to school* | 8 | 4.2 |
| **Years of experience** |  |  |
| *<1 year* | 16 | 8.3 |
| *1 – 3 years* | 50 | 26.0 |
| *3 – 5 years* | 27 | 14.1 |
| *>5 years* | 99 | 51.6 |
| **Received training on meat safety** |  |  |
| *Yes* | 46 | 24 |
| *No* | 146 | 76 |
| **Is the butchery the only source of income?** |  |  |
| *Yes* | 131 | 68.2 |
| *No* | 61 | 31.8 |
| **Valid medical health certificate** |  |  |
| *Yes* | 49 | 25.5 |
| *No* | 143 | 74.5 |

### Butcher shop attributes

Most butcher shops (84.4%) sourced their meat from Dagoretti slaughterhouse, with smaller proportions obtaining meat from Njiru (9.9%), Burma market (5.2%), and Limuru (2.08%). Meat was primarily transported using designated meat transport vehicles (82.3%) or motorcycles (31.3%), with (13.5%) of shops using both methods. The median transportation time for carcasses from the slaughterhouse to the butcher shops was 60 minutes, though this varied by location: 60 minutes in Kawangware, Kangemi, and Huruma, and 30 minutes in Waithaka. Upon arrival at all butcher shops (100%), the first step was weighing and hanging the meat on hooks at an ambient temperature. Only 9.9% of butchers removed waste (inedible portions) after hanging the meat. As expected, all butcher shops sold beef meat (an inclusion criterion for the study), while 32.8% (n = 63) also sold goat meat and 4.7% (n = 9) sold mutton (Table 2).

**Table 2:** Characteristics of meat sold in butcher shops (May to October 2022)

| Meat type | Current Selling price<br>(Ksh) (Median) | Quantity Ordered<br>(Kgs) (Median) | Number of days the<br>meat is sold (Median) |
| --- | --- | --- | --- |
| Beef | 560 | 45 | 2 |
| Goat | 650 | 15 | 2 |
| Mutton | 680 | 24 | 2 |

Meat was typically sold over a two-day period. In most shops (59.4%), leftover meat was left hanging at room temperature on the first day and transferred to the freezer on the second day. In contrast, 40.6% reported storing unsold meat in the freezer at the end of each day. Additionally, more than one-third of shops (37.5%; n = 72) also sold ready to eat foods including roasted meat (73.6%), stewed meat (72.2%), soup (29.2%), and *mutura* (African sausage) (16.7%).

For fly control, 58.3% of butcher shops used chemicals repellants, most commonly the fly-repellent coil *Baoma*. Other measures included maintaining cleanliness (28.7%), using a fly whisk (25.1%), traps (2.1%), and alternative methods such as diesel oil or candles (3.1%). One shop reported no action against flies, while another stated that flies were not present. For pest and rodent control, 46.8% of shops used chemical products, 14.6% relied on traps, and 44.8% employed other methods, including cats (7%) and cleanliness (9.3%). Notably, 35.9% of butcher shops reported no presence of rodents or pests.

### Food safety knowledge

Butchers demonstrated good knowledge in several areas, including the importance of government inspection and approval of meat before sale (98.4%), and checking freezers to ensure they are working properly (95.8%). Many also acknowledged the need to separate offal from other meat during transportation, processing, and storage (87%), and to avoid selling meat from an animal recently treated with antibiotics (83.9%). A detailed breakdown of responses to individual knowledge items is provided in Table 3.

**Table 3:**
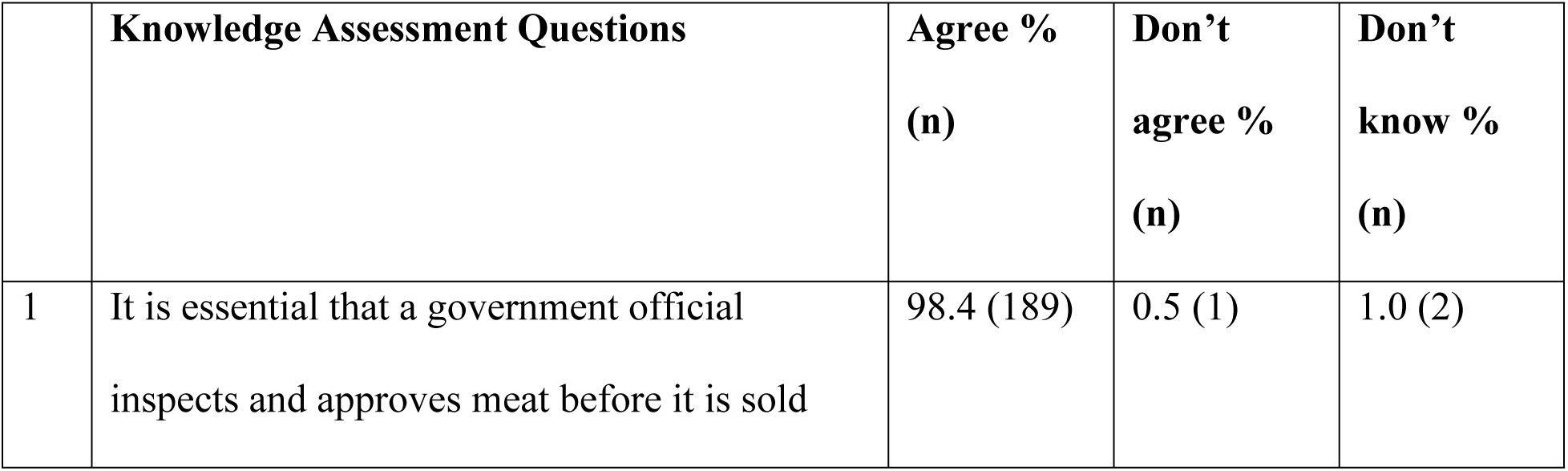
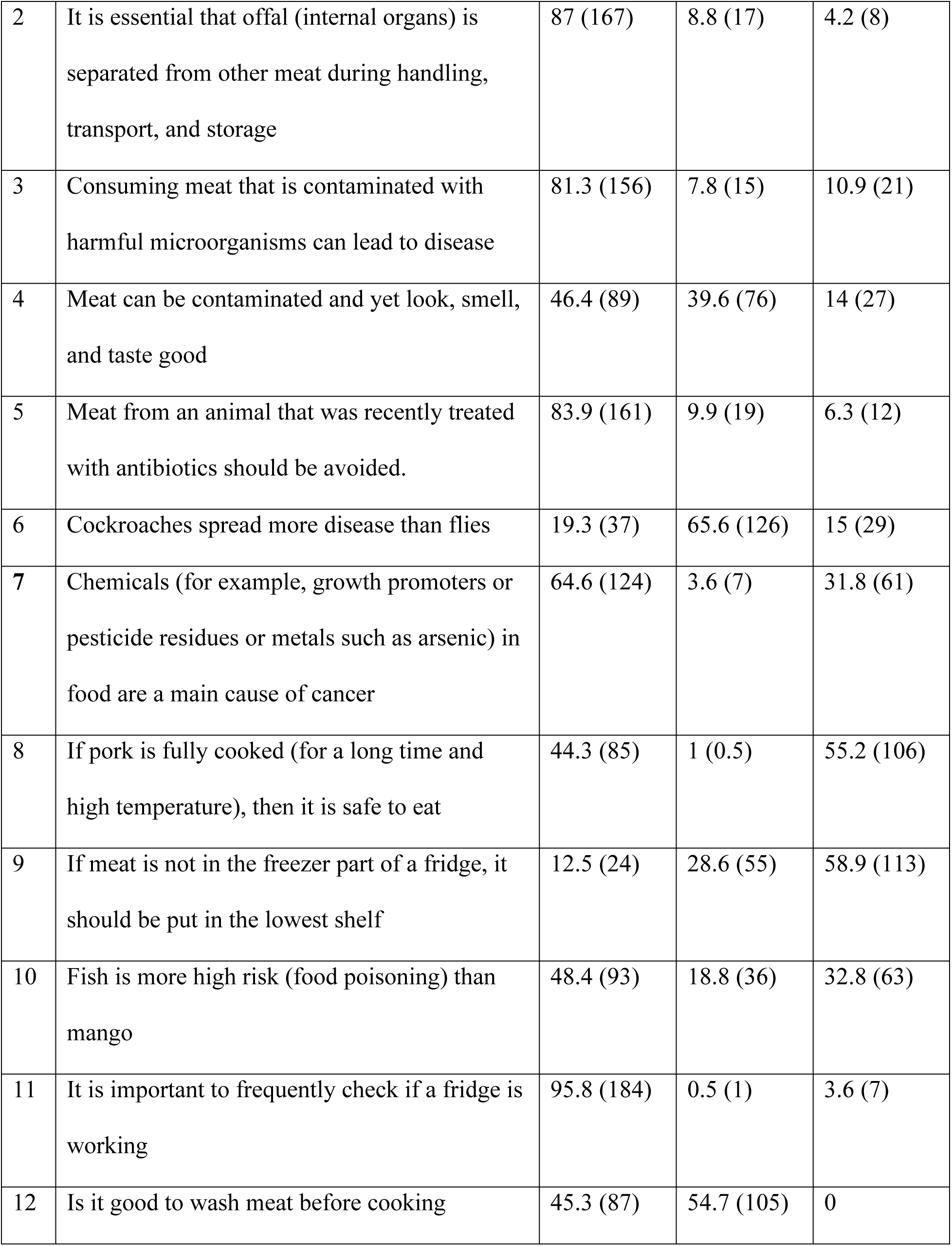
Butchers Food Safety Knowledge Responses.

|  | **Knowledge Assessment Questions** | **Agree %**<br>**(n)** | **Don't**<br>**agree %**<br>**(n)** | **Don't**<br>**know %**<br>**(n)** |
| --- | --- | --- | --- | --- |
| 1 | It is essential that a government official inspects and approves meat before it is sold | 98.4 (189) | 0.5 (1) | 1.0 (2) |
| 2 | It is essential that offal (internal organs) is separated from other meat during handling, transport, and storage | 87 (167) | 8.8 (17) | 4.2 (8) |
| 3 | Consuming meat that is contaminated with harmful microorganisms can lead to disease | 81.3 (156) | 7.8 (15) | 10.9 (21) |
| 4 | Meat can be contaminated and yet look, smell, and taste good | 46.4 (89) | 39.6 (76) | 14 (27) |
| 5 | Meat from an animal that was recently treated with antibiotics should be avoided. | 83.9 (161) | 9.9 (19) | 6.3 (12) |
| 6 | Cockroaches spread more disease than flies | 19.3 (37) | 65.6 (126) | 15 (29) |
| 7 | Chemicals (for example, growth promoters or pesticide residues or metals such as arsenic) in food are a main cause of cancer | 64.6 (124) | 3.6 (7) | 31.8 (61) |
| 8 | If pork is fully cooked (for a long time and high temperature), then it is safe to eat | 44.3 (85) | 1 (0.5) | 55.2 (106) |
| 9 | If meat is not in the freezer part of a fridge, it should be put in the lowest shelf | 12.5 (24) | 28.6 (55) | 58.9 (113) |
| 10 | Fish is more high risk (food poisoning) than mango | 48.4 (93) | 18.8 (36) | 32.8 (63) |
| 11 | It is important to frequently check if a fridge is working | 95.8 (184) | 0.5 (1) | 3.6 (7) |
| 12 | Is it good to wash meat before cooking | 45.3 (87) | 54.7 (105) | 0 |

A high proportion of participants provided incorrect responses to key food safety questions. For instance, 12.5% incorrectly identified where meat should be stored in a fridge, and 65.5% mistakenly believed that cockroaches spread more diseases than flies. Additionally, 54.7% wrongly thought washing meat before cooking was beneficial, and 44.3% incorrectly stated that sufficient cooking is enough to ensure food safety. Furthermore, 64.6% erroneously believed that chemicals in food are a major cause of cancer. When asked about awareness of the WHO Five Keys to Food Safety and HACCP, nearly all participants (99%) reported having no prior knowledge of these concepts. Additionally, visual tests of food safety knowledge revealed similar deficiencies. When shown five images depicting potential cross-contamination scenarios, participants frequently misclassified them, particular those involving sealed products where contamination was unlikely (Table 4).

**Table 4:**
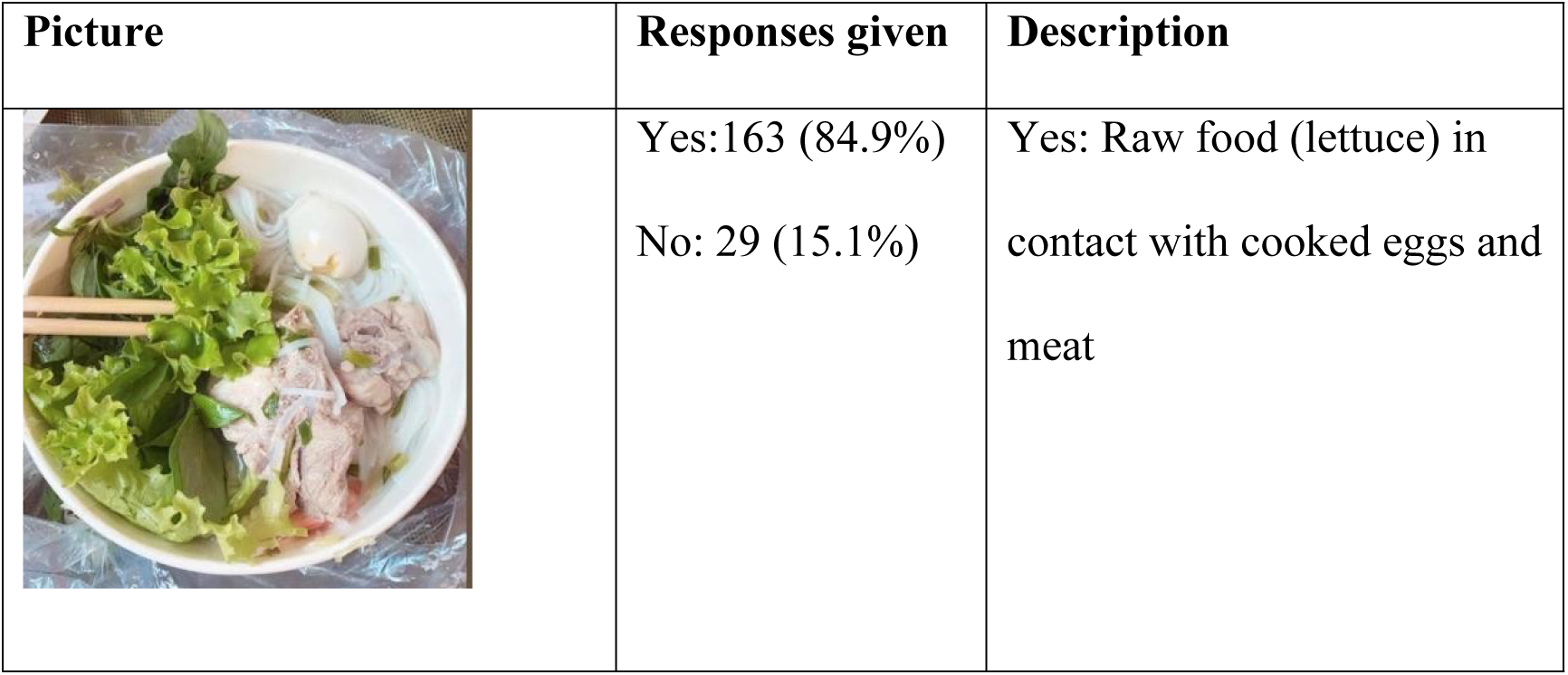
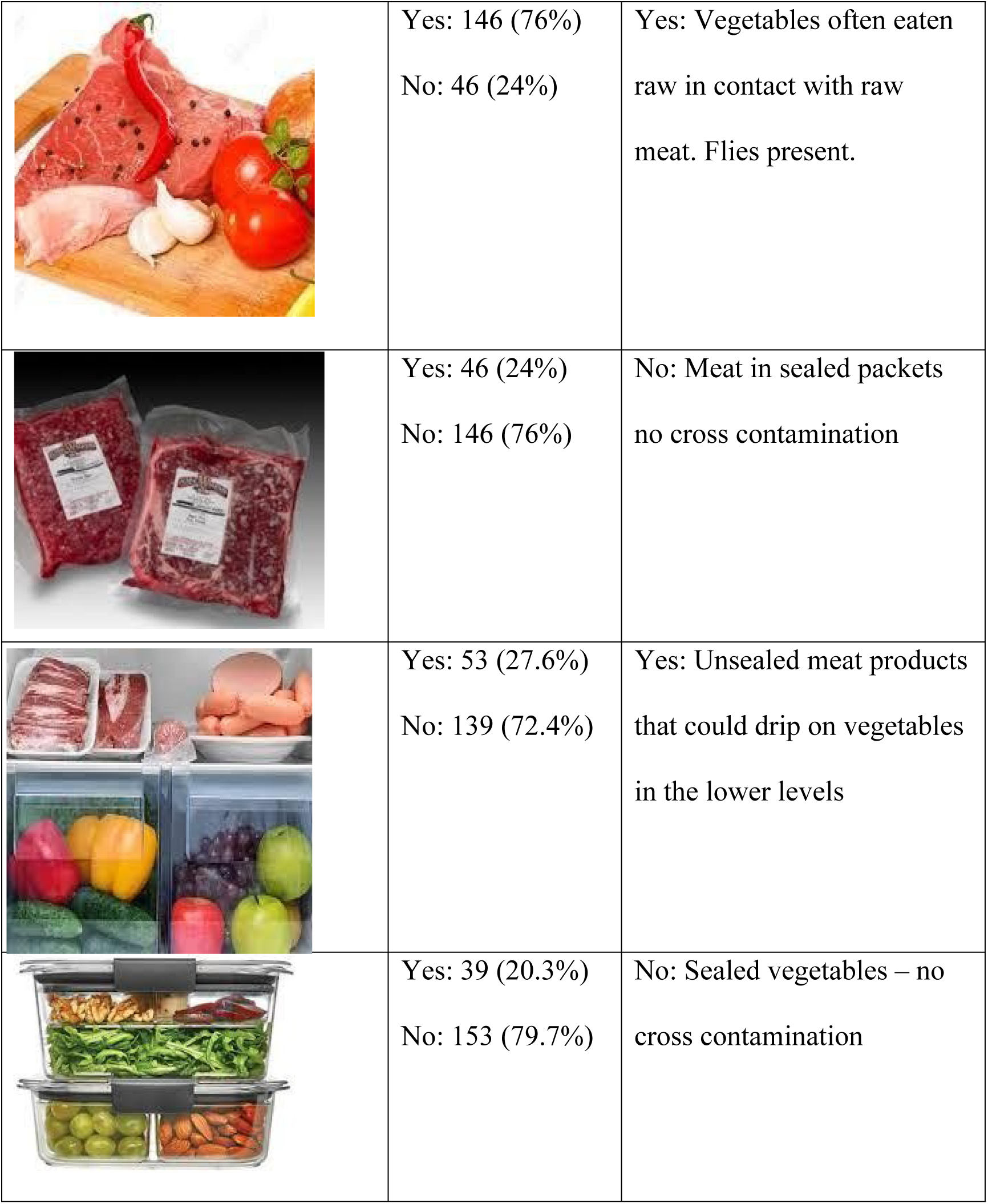
Food Safety Knowledge Images responses.

| Picture | Responses given | Description |
| --- | --- | --- |
|  | Yes: 163 (84.9%)<br>No: 29 (15.1%) | Yes: Raw food (lettuce) in contact with cooked eggs and meat |
|  | Yes: 146 (76%)<br>No: 46 (24%) | Yes: Vegetables often eaten raw in contact with raw meat. Flies present. |
|  | Yes: 46 (24%)<br>No: 146 (76%) | No: Meat in sealed packets no cross contamination |
|  | Yes: 53 (27.6%)<br>No: 139 (72.4%) | Yes: Unsealed meat products that could drip on vegetables in the lower levels |
|  | Yes: 39 (20.3%)<br>No: 153 (79.7%) | No: Sealed vegetables – no cross contamination |

### Food safety attitude

Butchers generally expressed positive attitudes to practices such as handwashing (96.4%), wearing protective clothing even in hot weather (69.8%), and avoiding work when sick (59.5%). However, less favourable attitudes were also common. Over 70% believed chilled meat loses flavor, face masks were uncomfortable (56.3%), and 53.6% did not regard foodborne diseases as a serious illness. In addition, 40.6% felt compelled to work even when sick due to a lack of alternatives. Attitudes towards authorities and regulation were mixed: while 52.1% viewed officials as allies, only 28.1% supported compliance with documentation requirements. A summary of the responses is shown in Table 5.

**Table 5:** Food safety attitude assessment of Butchers’ (May to October 2022)

|  | **Attitude Assessment Questions** | **Agree %**<br>**(n)** | **Don't agree**<br>**% (n)** | **Don't know**<br>**% (n)** |
| --- | --- | --- | --- | --- |
| 1 | Hand washing is enjoyable | 96.4 (185) | 3.1 (6) | 0.5 (1) |
| 2 | Protective clothing is too uncomfortable in hot weather | 29.7 (57) | 69.8 (134) | 0.5 (1) |
| 3 | You learn more by doing something than studying in class | 79.7 (153) | 16.7 (32) | 3.6 (7) |
| 4 | The authorities are butcher's friend | 52.1 (100) | 43.8 (84) | 4.2 (8) |
| 5 | In this economy, you have to work even if you are feeling sick | 40.6 (78) | 59.4 (114) | 0 |
| 6 | Chilled meat loses flavor | 72.4 (139) | 21.9 (42) | 5.7 (11) |
| 7 | Face masks are comfortable to wear | 43.8 (84) | 56.3 (108) | 0 |
| 8 | When it comes to meat, what you pay is what you get | 94.8 (182) | 4.7 (9) | 0.5 (1) |
| 9 | Foodborne disease is usually not a serious illness | 53.6 (103) | 37.5 (72) | 8.9 (17) |
| 10 | I can run this business without the required documents | 28.1 (54) | 69.8 (134) | 2.1 (4) |

### Food safety practices

Good food safety practices were observed in some domains. All reported buying meat inspected by government officials. Most washed or replaced towels daily (81.8%) and replaced aprons daily (74.5%). However, compliance was much lower for other requirements: fewer than half (42.7%), had all necessary approvals for handling and processing, only 1% stored chilled meat, and 6more than two-thirds (68.2%) ate at their workstations. When it came to handwashing, 61.1% of butchers indicated that they always washed their hands after coughing and sneezing. A summary of the various food safety practices is summarized in figure 2.

**Figure 2:**
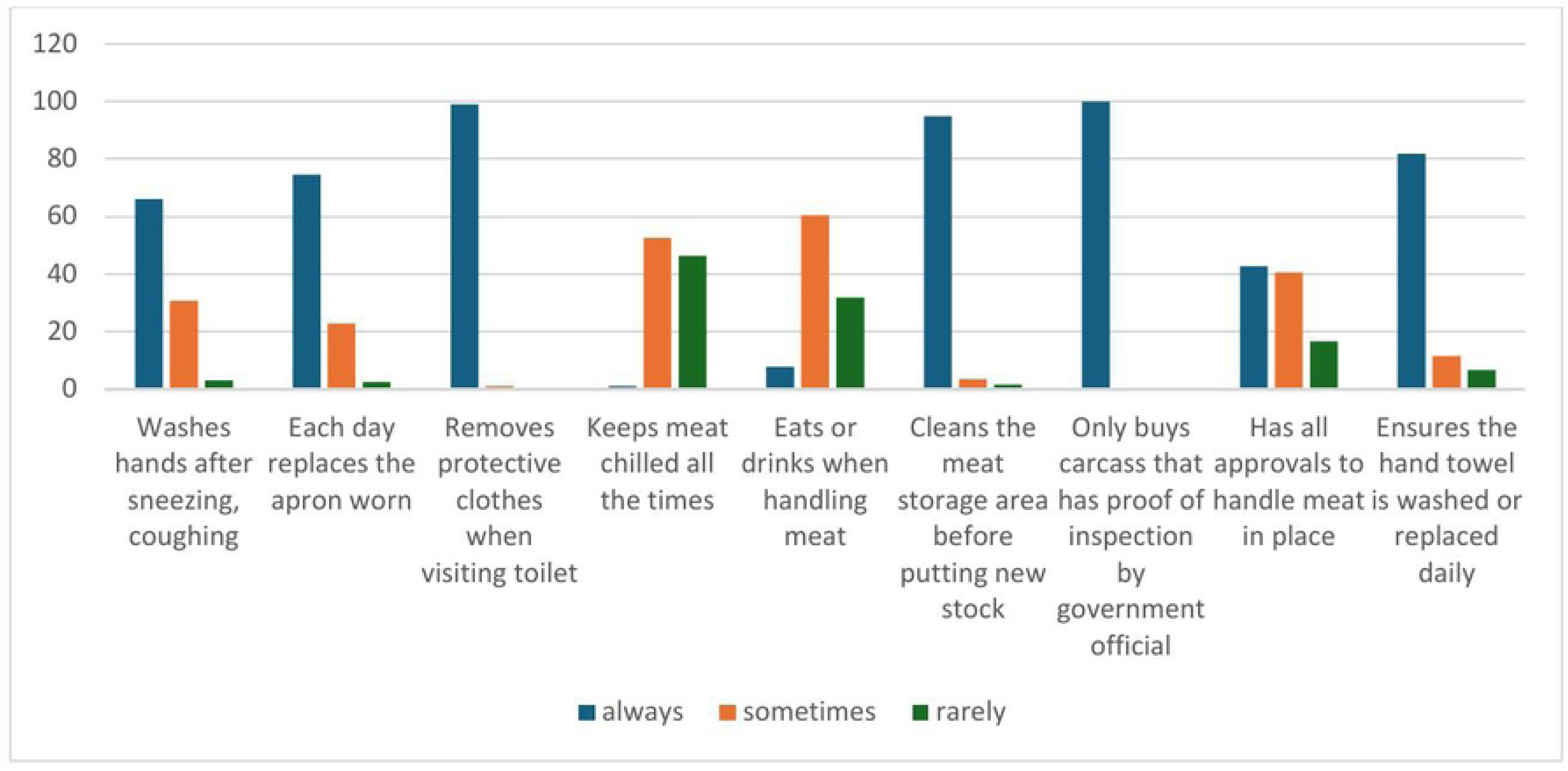
Practice assessment responses of butchers

When presented with images of poor practices, a large proportion admitted engaging in them: washing chicken carcasses under a tap (68.2%), cleaning blood with reusable cloths (71.4%), or wiping hands on dirty aprons (89.6%) (Table 6).

**Table 6:**
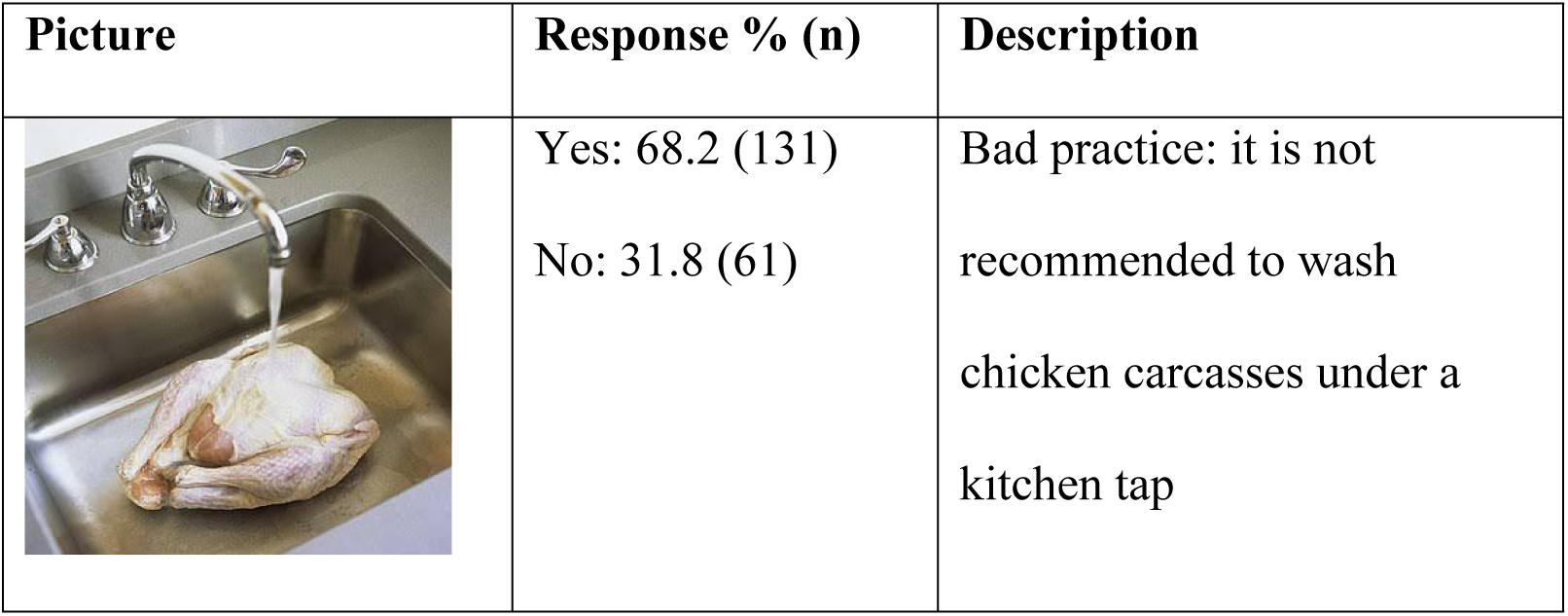
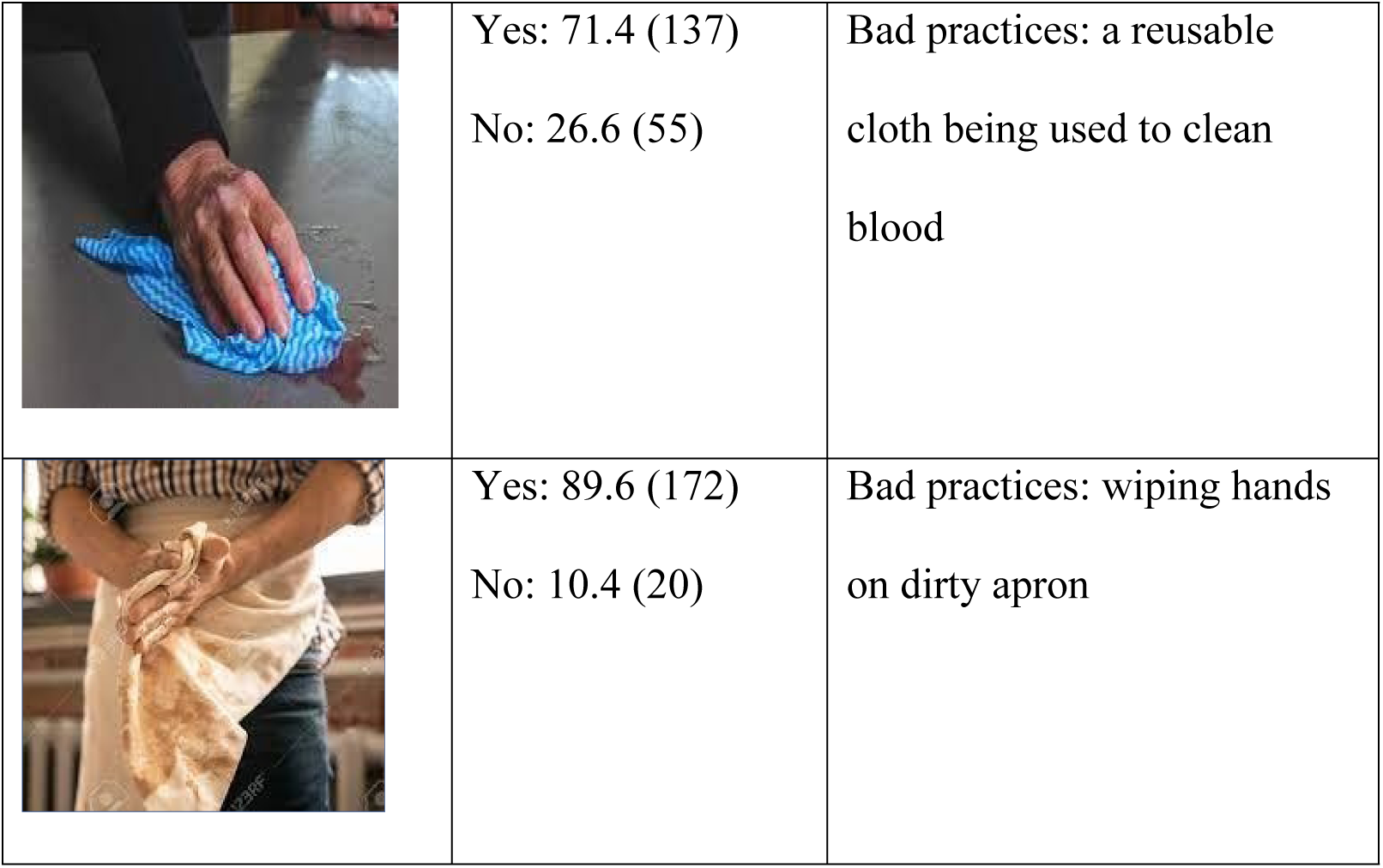
Food Safety Practice Image Assessment.

| Picture | Response % (n) | Description |
| --- | --- | --- |
|  | Yes: 68.2 (131)<br>No: 31.8 (61) | Bad practice: it is not recommended to wash chicken carcasses under a kitchen tap |
|  | Yes: 71.4 (137)<br>No: 26.6 (55) | Bad practices: a reusable cloth being used to clean blood |
|  | Yes: 89.6 (172)<br>No: 10.4 (20) | Bad practices: wiping hands on dirty apron |

Cleaning routines varied by surface. Most reported cleaning knives, cutting boards, weighing scales, and floors daily (>98%), but cleaning of hooks, meat containers, and walls was less frequent, often weekly or monthly (Table 7).

**Table 7:** Number of butchers that performed the various cleaning tasks.

| Cleaning task | Daily | Weekly | Monthly | Rarely |
| --- | --- | --- | --- | --- |
| **Cutting board** | 189 | 3 | 0 | 0 |
| **Weighing scale** | 188 | 3 | 0 | 1 |
| **Knife** | 191 | 1 | 0 | 0 |
| **Hanging hooks** | 119 | 67 | 1 | 5 |
| **Floor** | 190 | 2 | 0 | 0 |
| **Meat container** | 168 | 22 | 0 | 2 |
| **Walls** | 73 | 79 | 17 | 23 |

### Psychometric validation of KAP indices

Psychometric analysis supported the utility of the KAP indices. The knowledge scale showed a unidimensional factor structure, with the first principal component accounting for most of the variance. Internal consistency was modest (Cronbach’s alpha = 0.53), but acceptable for exploratory work (as in the case of our study). Item loading varied, with some negative or low correlations indicating heterogeneous item behavior. Further IRT analysis indicated that the scale was most informative for identifying butchers with lower knowledge levels, as information curves peaked below the mean theta. The attitude and practice scales demonstrated adequate psychometric performance under graded response models. Most items had moderate to high discrimination (>0.5) and well-ordered threshold parameters. Correlations among the latent trait scores for knowledge, attitude, and practice were modest (r = 0.16–0.27), suggesting that the three domains capture related but distinct dimensions of food safety behavior.

### Assessing association between KAP and demographic variables

No significant associations were observed between KAP indices and demographic variables such as education level, gender, work experience, age, or prior training.

### Level of bacterial contamination present in the hands of the butchers

Handprints cultures taken before and after meat handling revealed widespread contamination. Prior to handling, 64% of butchers’ hands showed heavy bacterial growth, 26% medium growth, and 9% low growth. After handling, heavy growth increased to 85.5%, with only 1.5% showing low growth. No associations were found between hand contamination and KAP indices (Table 8).

**Table 8:** Level of bacterial contamination in the hands of butchers.

|  | **Handprint One n (%)** | **Handprint Two n (%)** |
| --- | --- | --- |
| **Heavy growth** | 128 (64%) | 171 (85.5%) |
| **Medium Growth** | 52 (26%) | 24 (12%) |
| **Low growth** | 18 (9%) | 3 (1.5%) |
| **No handprint** | 2 (1%) | 2 (1%) |

### Intervention preferences

Seven potential interventions were proposed to butchers (Tables 9-10). Government-led food safety campaigns showed overwhelming support, with over three-quarters of butchers (78.1%) selecting this as their most preferred intervention. This was followed by cooperative group approaches (40.6%) and tamper-proof traceability labels (30.7%). In contrast, market-driven interventions faced considerable resistance. Hand-held freshness testing devices were the most rejected intervention, with 57.8% of butchers selecting this as their least preferred option and being chosen as “best” by only 14.1%. Similarly, high-quality brand labels were rejected by nearly half of respondents (49.5%) while receiving minimal endorsement (9.9%). Third-party testing by hospitals or universities with publication of results also faced substantial opposition (41.7% rejection), suggesting resistance to external oversight mechanisms.

**Table 9:** Interventions considered “best” by the butchers.

| **Intervention** | **% (n=192)** |
| --- | --- |
| Campaign for food safety & promote meat consumption | 78.13 |
| Co-operative group agreeing to follow good practices | 40.63 |
| Tamper-proof label showing farm, abattoir and shop | 30.73 |
| A certification scheme so trained butchers get a certificate | 16.67 |
| A hand-held device that measures meat freshness | 14.06 |
| Weekly meat testing by third party who makes results public | 9.90 |
| A brand showing meat is high quality | 9.90 |

**Table 10:** The intervention considered “worst” by the butchers.

| **Intervention** | **% (n=192)** |
| --- | --- |
| A hand-held device that measures meat freshness | 57.81 |
| A brand showing meat is high quality | 49.48 |
| Weekly meat testing by third party who makes results public | 41.67 |
| Tamper-proof label showing farm, abattoir and shop | 18.23 |
| A certification scheme so trained butchers get a certificate | 14.58 |
| Campaign for food safety & promote meat consumption | 9.38 |
| Co-operative group agreeing to follow good practices | 8.85 |

### Latent characteristics of butchers

The initial unrotated solution indicated that three factors had eigenvalues > 1, together accounted for 69.8% of the total variance (Factor 1 = 27.5%; Factor 2 = 21.6%; Factor 3 = 20.6%). The rotated factor loadings revealed a coherent and interpretable structure (Table 10), with each factor representing a distinct preference dimension:

- **Factor 1: Support-Seeking, Collectivist versus Market-Based Interventions** This factor showed strong positive loadings for group-based and government campaign interventions and strong negative loadings for brand label and hand-held device. It represents a dimension contrasting a preference for supportive, familiar, and collective interventions with rejection of market-driven or technological mechanisms that may be perceived as judgmental or consumer-empowering.
- **Factor 2: Embracing Formal Schemes and Traceability** This factor was defined by strong positive loadings for certification schemes and traceability labels (which in Kenya are both typically government schemes), indicating a latent preference for formalized, often government-or authority-led quality assurance schemes.
- **Factor 3: Disliking Independent, Effective Oversight** The third factor was characterized by a strong negative loading on third-party testing and a moderate positive loading on government campaign. This pattern suggests a distinct concern or discomfort with external, possibly more rigorous scrutiny by trusted non-governmental institutions, such as universities or hospitals. In contrast, government campaigns may be perceived as less threatening or more lenient forms of oversight.

Loadings and uniqueness values are presented in Table 11. Most variables exhibited moderate to strong communalities, with uniqueness values below 0.40, confirming that the retained factors explained a substantial portion of variance.

**Table 11:**
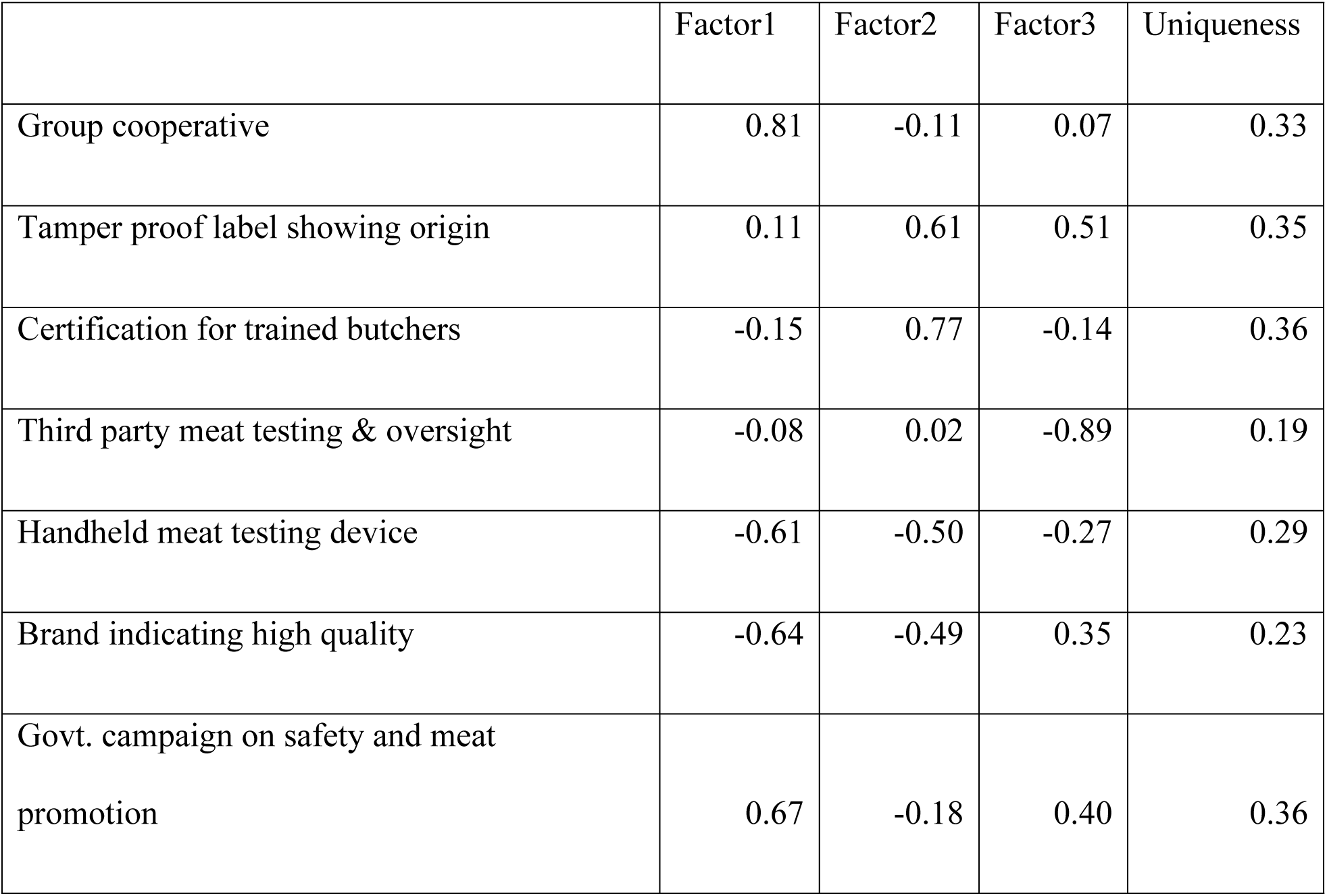
Loadings and uniqueness values for proposed food safety interventions.

Handheld devices load negatively on both Factor 1 (−0.61) and Factor 2 (−0.50), suggesting rejection by butchers who favored collective approaches or formal systems. Interesting, the minority who preferred handheld testing devices scored significantly higher on knowledge (5.6 vs 4.9, p=0.04), attitudes (5.8 vs 4.9, p=0.009) and practices (5.5 vs 4.9, p=0.08). Conversely, those favouring cooperatives had significantly lower knowledge (4.7 vs 5.2 p=0.03) and practice scores (4.7 vs 5.2, p=0.04) and tended to score lower on attitude (4.9 vs 5, not significant).

### Butcher typologies

The Calinski-Harabasz criterion suggested that a three-cluster solution provided optimal separation (pseudo-F = 73.97), which was supported by dendrogram inspection (Figure 3).

**Figure 3:**
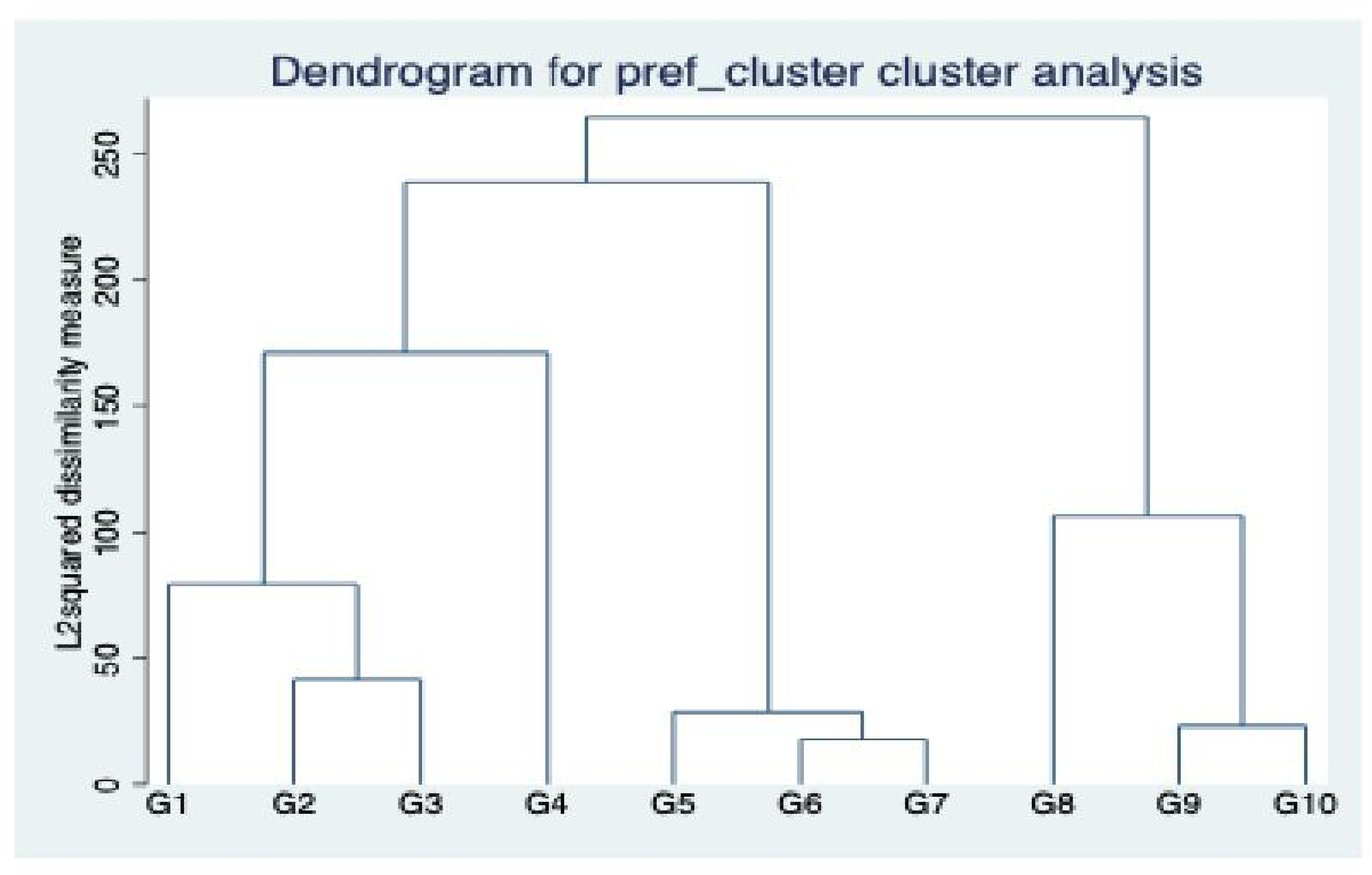
Dendrogram showing clustering of butchers according to KAP

The three clusters differed significantly in their mean factor scores (all p<0.001) and represented distinct butcher typologies:

Cluster 1: “Collectivist Effective Oversight-Avoiders” (n=92, 47.9%) exhibited near-neutral preferences for collective versus individual approaches (f1 = 0.01), slight aversion to formal certification systems (f2 = −0.33), but strong avoidance of external oversight (f3 = 0.75). This largest group preferred supportive interventions while strongly rejecting rigorous external scrutiny.

Cluster 2: “Forward-Facing Self-Reliant” (n=29, 15.1%) showed strong preference for individual-based interventions (f1 = −1.71), slight aversion to formal systems (f2 = −0.25), but willingness to accept external oversight (f3 = −0.82). This smallest group appeared confident in their practices and open to objective measurement tools.

Cluster 3: “Formal System-Embracers” (n=71, 37.0%) demonstrated preference for collective approaches (f1 = 0.69), strong endorsement of formal certification systems (f2 = 0.53), and acceptance of external oversight (f3 = −0.64). This group favored official, institutionalized quality assurance mechanisms.

Statistical validation confirmed the robustness of the three-cluster solution. ANOVA revealed highly significant differences across all three factor scores (F₁ = 155.53, F₂ = 19.27, F₃ = 104.80; all p < 0.001), indicating excellent cluster separation. Cross-validation with intervention preferences reinforced the interpretation: Forward-Facing Self-Reliant group significantly preferred handheld testing devices (< 0.001) and third-party testing (p < 0.001), while Collectivist Oversight-Avoiders and Formal System-Embracers strongly favored government campaigns (p < 0.001).

## Discussion

Demographic characteristics influence food safety risks and inform the planning of interventions. In this study, most butchers were male, a finding consistent with previous studies in East and West Africa [20, 21, 22, 23, 24, 25]. Most butchers were aged between 21-30 years, younger than findings from other studies, which commonly reported meat handlers aged 31-40 years [22, 24, 26, 27, 28]. Half of the butchers had secondary education, consistent with a study in Kenya [21] but contrasting with other studies that reported primary education as the norm [22, 24, 26, 29]. Most butchers (66.7%) were employees rather than shop owners, a finding contrary to studies in Ghana [25] and Ethiopia [22] where shop owners dominate. Many butchers had more than five years of experience handling meat, aligning with previous studies [22, 25, 26]. Lastly, despite being required to have a medical health certificate, a prerequisite in the [30], many of the butchers did not have the medical health certificate, a finding similar to other studies [20, 22, 24, 28].

While butchers demonstrated adequate knowledge of certain food safety principles, significant gaps were evident. For instance, nearly all were unfamiliar with the WHO’s Five Keys to Food Safety or HACCP. Moreover, most butchers lacked food safety training, similar to findings in Ethiopia [22, 24] and Kenya [20, 21]. Training is essential, as studies show that it significantly improves food safety KAP [31] Good knowledge was noted regarding practices like separating offal from other meat and monitoring freezer functionality. However, knowledge of hygiene— such as contamination risks, pest control, and meat storage—was weaker, despite its critical importance. Observations suggest that the presence of veterinary officers inspecting meat may explain higher compliance awareness.

There was no correlation between the demographic features of butchers (age, level of education, gender, years of experience, and training) and their degree of food safety knowledge in this study. Another study [32] also reported no associations between gender, level of education, and years of experience, and the level of food safety knowledge. However, others have found significant associations, as theory would suggest [26].

Food safety attitude is an important element as it can guide food safety practices among butchers. While good attitude was shown to some hygiene practices, poor attitude was shown regarding the severity of foodborne infections, chilled meat, working while sick, and the relationship between butcher shops and the authorities. While most of the butchers responded that they could not run the business without the required documents, it was observed that a large percentage did not in actuality have the required documents to run the butcher shops. There was no association between the butchers’ demographic factors and food safety attitude. This was similar to the study [32] that also reported no association between socio-demographic factors (gender, tribe, religion, level of education, marital status, age, and years of experience) and food safety attitude.

Food safety practices directly influence the quality of meat sold in retail enterprises. In this study, hand washing and PPE wearing behaviors were higher than reported in similar studies in Ethiopia and Nigeria [22, 24, 33]. There was no association between the butchers’ demographic features (gender, age, level of education, working experience, and training) and the level of food safety practices. This was contrary to a study by [32] which reported an association between level of education, marital status, age, years of experience, and tribe.

Although knowledge was reasonable and attitudes generally positive most butchers had a heavy growth level of bacterial contamination in their hands with an increase in bacterial contamination on their hands after handling and cutting meat. This contamination can be due to poor hand washing practices and personal hygiene. During data collection, it was observed that most butcher shops did not have hand washing stations and used the same cloth towel to wipe surfaces, equipment and hands after meat handling. Very high levels of meat contamination have been previously reported in Kenya [15, 20, 34], and people have died after consuming contaminated meat [35]. Our findings on adequate KAP yet high contamination align with emerging evidence that food safety behaviour in LMICs is shaped by a complex interplay of knowledge, motivation, opportunity, and structural constraints and requires behaviorally informed interventions that go beyond traditional knowledge-based training to address both psychological drivers and enabling conditions, as framed by the COM-B and other behavior change models [36].

Unexpectedly, there was no association between demographic factors, the level of food safety knowledge and practice and the level of microbial hand contamination. While others have found this [37], theory would suggest an association, and this is also reported [38]. The food safety knowledge and practice gap is real. Several studies have shown that good or sufficient knowledge does not reflect or translate to good practice [24,33]. However, enabling conditions and structural constraints dominate informal markets where meat is sold [39]. Many shops lacked handwashing facilities, had inadequate water and sanitation – with most shops relying on buying water from third party vendors using buckets. We also observed weak enforcement of existing laws as most butchers lacked the required medical certificate and mandatory food safety training. Additionally, the adoption and use of cold chain is deterred by unreliable power, high electricity bills, and customer preference of meat that is not frozen or kept in the fridge. The butchers reported that most consumers claim that frozen meat loses its flavor. The butchers also claimed lack of incentives or motivation to promote better food safety practices, similar to another study [39]. It is essential to note that most of the butcher shop locations were wanting, with most surrounded by open running sewage.

In our study, contamination was much higher than anticipated (64% at maximum level before meat handling and 86% at maximum after handling. The lack of association between butcher’ KAP and hand contamination may be as a result of a ceiling effect which reduced our ability to make distinctions between butchers and reveal associations between KAP and microbial outcomes. However, it may also suggest structural determinants such as regulatory systems, infrastructure, and market incentives which play a vital role in shaping food safety KAP and microbial contamination than individual characteristics. While carrying out the study in the field, we noticed that the butchers used the same towel to wipe their hands after handling meat and to clean the butcher shop surfaces, knives and weighing scale, an action that may explain the increase in hand microbial contamination.

Factor analysis revealed three distinct orientations toward support-seeking versus individual accountability, formal versus informal systems, and government versus independent oversight. The strong preference for government campaigns and cooperative groups aligns with Kenya’s cultural tradition of *harambee* (collective action), where community-based approaches to development are deeply embedded in social values. Conversely, the rejection of handheld testing devices and brand labels may reflect resistance to interventions that shift power toward consumers or create competitive advantages for individual butchers, potentially disrupting established market relationships.

The emergence of three distinct butcher typologies has important implications for intervention targeting. Rather than assuming uniform adoption of “evidence-based” interventions, our findings suggest that preference heterogeneity requires differentiated approaches. The “Forward-Facing Self-Reliant” group, though smallest (15%), represents butchers willing to embrace objective measurement tools, third party oversight, and quality branding suggesting they could serve as early adopters for technology-based and market-led interventions. Conversely, the majority “Collectivist Effective Oversight-Avoiders” may require supportive, non-threatening approaches that preserve existing social structures. Butchers preferring consumer-led and technology interventions had higher KAP than others while those preferring collectivist and government-led approaches had lower KAP congruent with their preferences for more innovative and modern approaches to food safety.

Although this large, probabilistic study gives key insights into KAP and intervention preferences it has some limitations. Practices could have been better assessed using an observation check list instead of a questionnaire to minimize self-reporting bias. Additionally, while bacterial contamination assessed from direct handprints provided a biological indicator of contamination, this method was not precise enough to fully explore the relationships between demographic characteristics, KAP and hand contamination, and using swabs or serial dilutions would have provided more information.

## Conclusion

The meat industry is a critical livelihood sector in Kenya, but food safety remains compromised by structural, behavioral, and culture barriers. Despite reasonable knowledge and generally positive attitudes, high bacterial contamination and poor hygiene practices were observed among butchers. Training and regulation alone are insufficient; interventions must address the enabling environment and underlying behavioral drivers. Conventional food safety interventions remain popular but have shown limited in practice. Most innovative approaches such as consumer-driven technologies and market-based incentives, face resistance but could be piloted with a minority of butchers who show readiness for change. These early adopters could serve as “change champions” in future food safety initiatives. Ultimately, effective improvement of meat safety in LMICs will require multi-pronged strategies: strengthening infrastructure and enforcement, embedding behavioral science into training, leveraging cultural traditions of collectivism, and selectively introducing new technologies through willing participants. By acknowledging heterogeneity in preferences and the structural realities of informal food systems, more context-appropriate and sustainable interventions can be designed.

## Data Availability

All relevant data are within the manuscript and its Supporting Information files.

## Author contributions

WO – conceptualization, data curation, formal analysis, investigation, methodology, software, validation, visualization, writing – original draft; PO: Writing – review & editing, Supervision; PK: Writing – review & editing, investigation, validation, data collection, data entry and cleaning; MK: writing-review and editing, analysis; MK: Writing – review & editing, Software; LO: Writing – review & editing, Supervision; AM: Writing – review & editing, Supervision, Conceptualization, Resources, Methodology; DG – conceptualization, funding acquisition, formal analysis, data curation, writing - review and editing; FM: Writing – review & editing, Supervision, Resources, Project administration, Methodology, Funding acquisition, Conceptualization. All authors read, commented and approved of the final manuscript.

## Acknowledgements

The authors thank Vétérinaires Sans Frontières who were involved the data collection. We are also grateful to the individual butchers who generously shared their knowledge and experience with us by participating in the study.

## Data Availability

The data supporting this study are accessible upon request from the authors.

